# Automated imaging-based tumor burden and pre-treatment circulating tumor DNA in HPV-associated oropharynx cancer

**DOI:** 10.1101/2025.01.15.25320598

**Authors:** Mina Bakhtiar, Zezhong Ye, Jonathan D. Schoenfeld, Homan Mohammadi, Jeffrey P. Guenette, Eleni M. Rettig, Glenn J. Hanna, Benjamin H. Kann

## Abstract

**Background:** Artificial intelligence (AI)-based imaging analysis has applications for the diagnosis of head and neck malignancies, and serum circulating tumor-associated DNA (ctDNA) is an emerging biomarker being evaluated for response assessment and risk stratification in human papilloma virus (HPV)-associated oropharynx squamous cell carcinoma (HPV-OPSCC). The relationship between automated imaging biomarkers and ctDNA has not yet been explored.

**Objective:** To test the association between ctDNA and AI-derived measures of tumor burden among patients with HPV-OPSCC.

**Design, Setting, and Participants:** This cross-sectional study included patients who were treated with curative intent for HPV-OPSCC between 2020-2023, prospectively enrolled on a blood collection protocol (Clinical trials.gov identifier: NCT04965792).

**Exposures:** Clinical factors including demographics, AJCC 8^th^ edition clinical staging, and HPV genotype.

**Main Outcomes and Measures:** Pre-treatment serum measurement of circulating tumor-tissue modified viral (TTMV) HPV-DNA using a commercially available test, measured as a continuous value (fragments/mL). Primary tumor and nodal volumes, total tumor volume, and cystic/necrotic nodal volume were generated on pre-treatment diagnostic or radiation CT- planning scans using a prospectively validated AI auto-segmentation algorithm. Assessments of model fit: Akaike information criterion (AIC) and Bayesian information criterion (BIC).

**Results:** 170 patients with HPV-OPSCC were included in the study. On univariable regression, primary tumor volume (coeff=39.43, p<0.001), nodal volume (coeff=39.54, p<0.001), AJCC 8^th^ edition Tumor (T) stage (coeff = 1031.09, p=0.009), Nodal (N) stage (coeff=1840, p=0.018), HPV subtype 16 (coeff=3072.40, p=0.006), and CCI (coeff=-596.60, p=0.038) were associated with ctDNA. Cystic nodal volume was not associated with ctDNA (coeff=0.31, p=0.11). On multivariable analysis, primary tumor and nodal volumes were associated with ctDNA (coeff=34.79, p=0.001 and coeff=24.68, p=0.022, respectively), but T and N stage were not (coeff=-439.28, p=0.37 and coeff=238.19, p=0.29, respectively). Including automated tumor and nodal volumes improved model fit compared to T and N stage alone (3420.96 vs 3435.88 AIC, 3449.18 vs 3457.83 BIC).

**Conclusions and Relevance:** AI-automated volumetrics on pretreatment imaging are independently associated with ctDNA, controlling for clinical stage. The association is stronger than staging and improved predictive capacity of regression models. AI-automated volumetrics may provide a practical correlate to ctDNA levels and help risk stratify patients.

## Introduction

There is a rising incidence of human papilloma virus (HPV)-driven oropharynx squamous cell carcinoma (HPV-OPSCC), with HPV-associated disease now comprising 70% of all newly diagnosed oropharynx cancers in the United States.^1^ While HPV-OPSCC is treatment responsive and often can be cured, there is growing interest in the use of liquid biomarkers for a personalized approach to management. One well-researched test is circulating tumor DNA (ctDNA) or the detection of tumor-shed DNA fragments in the blood. The drivers of circulating DNA levels in patients with HPV-OPSCC are still largely unknown, and the kinetics of ctDNA rise and clearance during disease progression and treatment are complex; understanding ctDNA variability and its relationship to disease burden is a rapidly growing area of interest. Existing studies demonstrate that the detection of tumor-associated HPV fragments has high diagnostic sensitivity and specificity^2–4^, and that pre-treatment measures of ctDNA have been associated with clinicopathologic features including lymphovascular invasion and clinical stage.^5^ Others highlight the role of ctDNA in surveillance, including detection of disease recurrence^6^ and in the assessment of metastatic disease burden.^7^ Additionally, patients who are cured of HPV-OPSCC may experience a high rate of short- and long-term treatment-related toxicity, and several prospective clinical trials have opened with the goal of treatment de-escalation (NCT03323463; NCT02281955; NCT03952585). Attempts to broadly de-intensify therapy for HPV-OPSCC have had variable success,^8^ and ctDNA is currently being evaluated as a method for improved risk-stratification (NCT04900623).

Despite these evolving clinical applications, there are limitations to the use of ctDNA, including time and cost of the assay.^9^ This has prompted investigation into the availability of a diagnostic complement to ctDNA. Imaging including head and neck CT or PET remains the standard for diagnosis and monitoring of HPV-OPSCC, and the relationship of ctDNA to imaging is an area of active investigation.

In this study, we set out to determine whether AI automated tumor and lymph node volumetrics derived from pre-treatment imaging were associated with ctDNA level. We investigated whether this association was stronger than, or independent of, AJCC 8^th^ edition clinical staging as a conventional measure of tumor burden. Finally, we sought to identify whether certain nodal characteristics like cystic or internally necrotic regions commonly seen in HPV-driven oropharynx cancer were associated with circulating DNA levels.

## Methods

This study was conducted in accordance with the Declaration of Helsinki guidelines and following Mass General Brigham Institutional Review Board approval. The study is reported in accordance with the TRIPOD (Transparent Reporting of a multivariable prediction model for Individual Prognosis or Diagnosis) guidelines. Waiver of informed consent was obtained. A secondary analysis was performed on a single-institution prospective cohort of patients (Clinicaltrials.gov identifier: NCT04965792) who were treated definitively for HPV-OPSCC and enrolled in a protocol for ctDNA-guided post-treatment surveillance.^10^ HPV-positivity was determined using p16 positivity on immunohistochemistry and/or positive for high-risk HPV by in situ hybridization or PCR. All patients included in the study had pre-treatment tumor tissue modified viral DNA drawn via a commercially available assay (TTMV, NavDX, Naveris) measured in the serum in fragments per ml. Clinical data was abstracted from electronic medical records, including patient demographics, disease characteristics, treatment details, and pre-treatment imaging, including diagnostic CT scans and PET-scans, or pre-radiation CT simulation scans. A subset of patients included in this study (N=79) were also included in a previously reported analysis of the association between ctDNA and pre-treatment clinicopathologic features.^5^

### Image Preprocessing

CT and PET images were first resampled to a spacing of 1×1×3 using linear interpolation on Python SimpleITK.^11^ This step was essential to improve generalization and achieve a more manageable size. Scans of the entire body were reduced to 65% of their original slice length to eliminate extraneous information from lower anatomical regions, such as the abdomen and pelvis. A fixed reference file was selected for rigid registration of all data files, ensuring coordinated correspondence across the scans. One scan was designated as the reference image, and all other data files were aligned to this template. Following registration, files were cropped to a resolution of 160×160×64 using a custom bounding box procedure, which was centered around the skull’s midpoint. To accurately identify the head region, we employed OpenCV^12^ to detect the largest elliptical contour in the upper two-thirds of the image, analyzing the area-to-perimeter ratios of all contours. We then calculated the centroid of the skull ellipse to center the crop. Finally, the CT scans were clipped using a Hounsfield window of [−200, 275] and normalized for consistency.

### Primary tumor and lymph node auto-segmentation

An AI auto-segmentation algorithm using a 3D nnU-Net backbone^13^ validated on a multi-institutional cohort^14^ was applied to segment tumor and malignant lymph nodes and to calculate tumor burden via 3D volumetrics on pre-treatment diagnostic or CT radiation planning scans. We calculated primary tumor volume, nodal volume, and total volume. Additionally, we calculated the volume of cystic or necrotic-appearing tissue within lymph nodes. Cystic or necrotic-appearing material was defined as the volume within lymph nodes with a density between –100 and +20 Hounsfield units (HU) similar to the range proposed in prior literature.^15^ Given there is no clearly defined HU threshold for cystic fluid, we also investigated −20 to +20 HU, consistent with water density. Cystic ratio was calculated as the volume of hypodense or necrotic fluid divided by the total lymph node volume (**Figure 1**).

**Figure 1.**
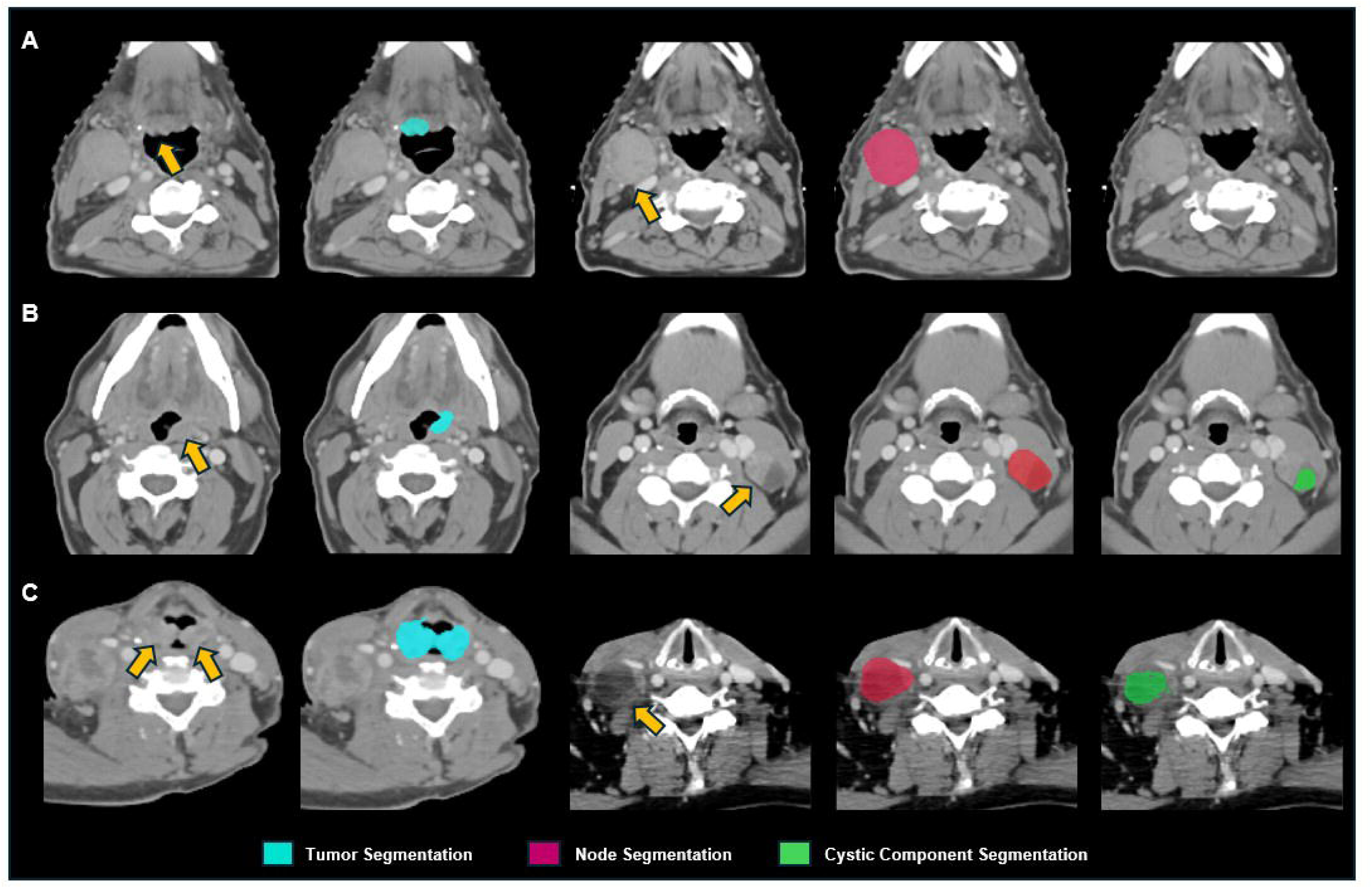
Sample Auto-segmentation of tumor, malignant lymph node and lymph node cystic component.

### Statistical Analysis

Descriptive statistics were generated to describe baseline patient demographics and disease characteristics. Spearman tests were performed to measure the correlation between ctDNA and automated volumes and the association between ctDNA and clinical stage. Single variable linear regression was performed to detect the association between ctDNA and automated volumes, AJCC 8^th^ edition clinical stage, and patient demographics and disease characteristics. Multivariable linear regression was performed to detect the association between ctDNA and automated volumes when controlling for smoking status (current, former, or never smoker), hpv subtype (16 vs other), AJCC 8^th^ edition tumor and nodal stage, and Charlson Comorbidity Index (CCI). Variables that met a significance threshold of p=0.05 in the single variable regression were included in the multivariable model. We compared model fit when including vs omitting automated tumor and node volumes using Akaike’s information criterion (AIC), Bayesian information criterion (BIC), and adjusted R^2^. All statistical analyses were conducted in Stata SE version 17 (StataCorp. 2021. *Stata Statistical Software: Release 17.* College Station, TX: StataCorp LLC).

## Results

Between 2020-2023, 170 patients with HPV-OPSCC were included in the study. Median age was 63.4 years (interquartile range [IQR] 57.9-69.5), 79% male. Eighty-two patients (48%) were never-smokers, 13 (8%) were current smokers and 75 (44%) were former smokers. One-hundred and sixty-six patients (98%) were treated for newly diagnosed HPV-OPSCC; 4 were treated for persistent or recurrent disease. The most common primary tumor site was the base of tongue (74 patients, 44%) followed by the tonsil (69 patients, 41%). HPV-16 was the most common subtype (130 patients, 76%), followed by HPV 33 (14 patients, 8%). One-hundred and ten patients (65%) were AJCC 8th edition clinical stage I, 34 (20%) were AJCC 8th edition clinical stage II, and 26 (15%) were AJCC 8th edition clinical stage III (**Table 1**). Mean TTMV was 2499.27 fragments/mL (Standard deviation: 6189.78). Twenty-one patients had a TTMV of 0 at diagnosis (**Table 2**, **Figures 2a-2d**).

**Figures 2a-2d.**
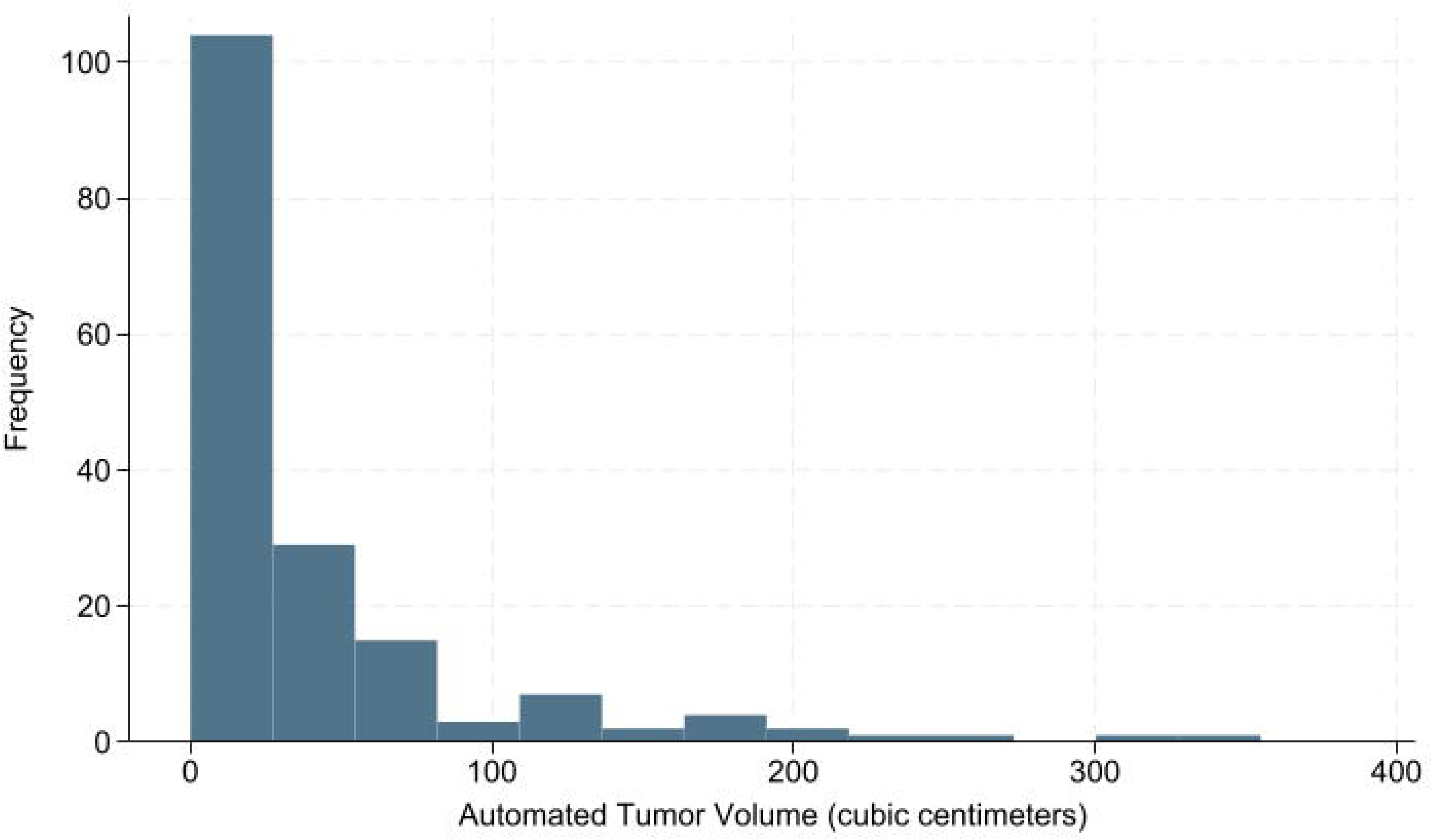
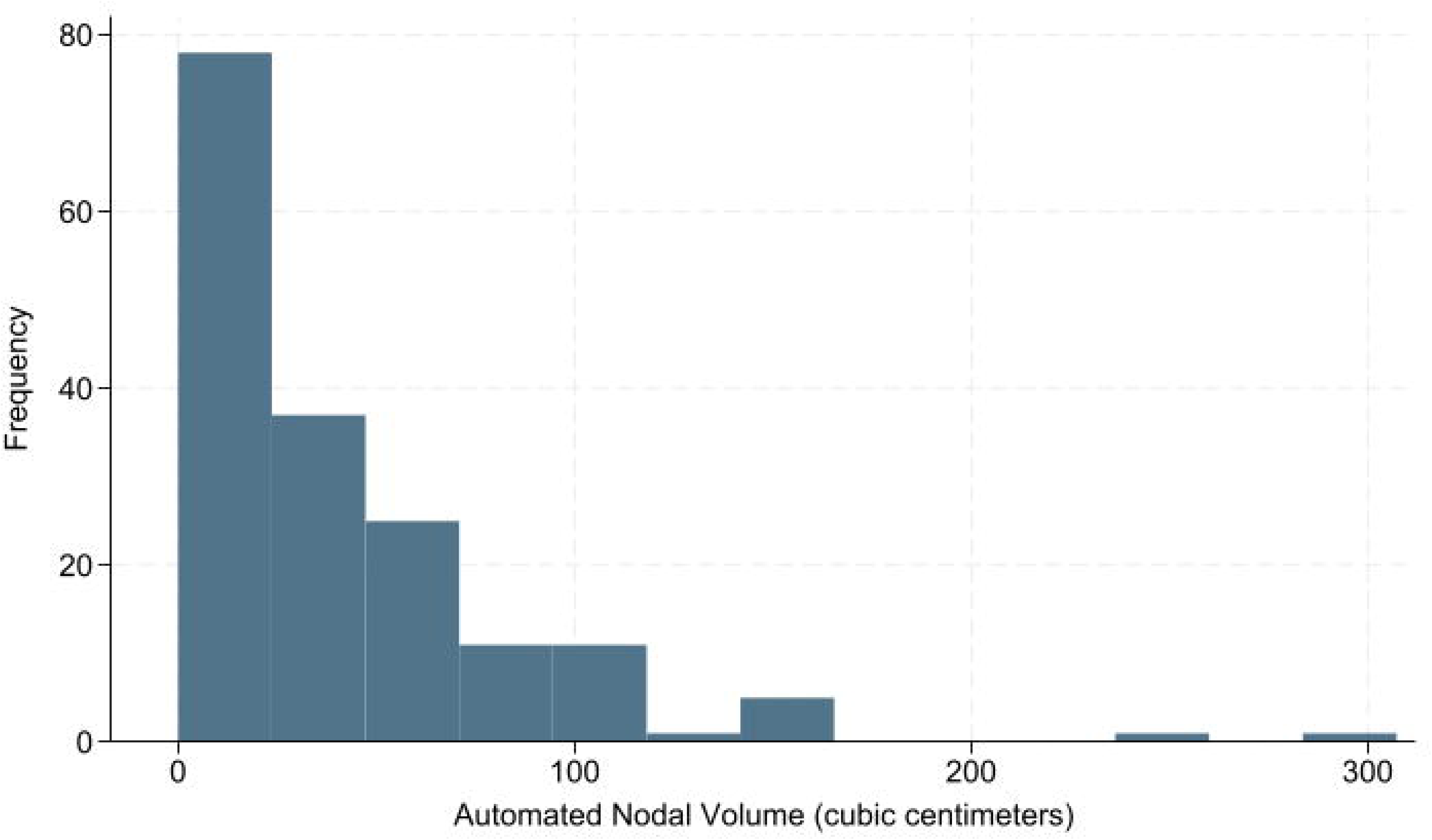
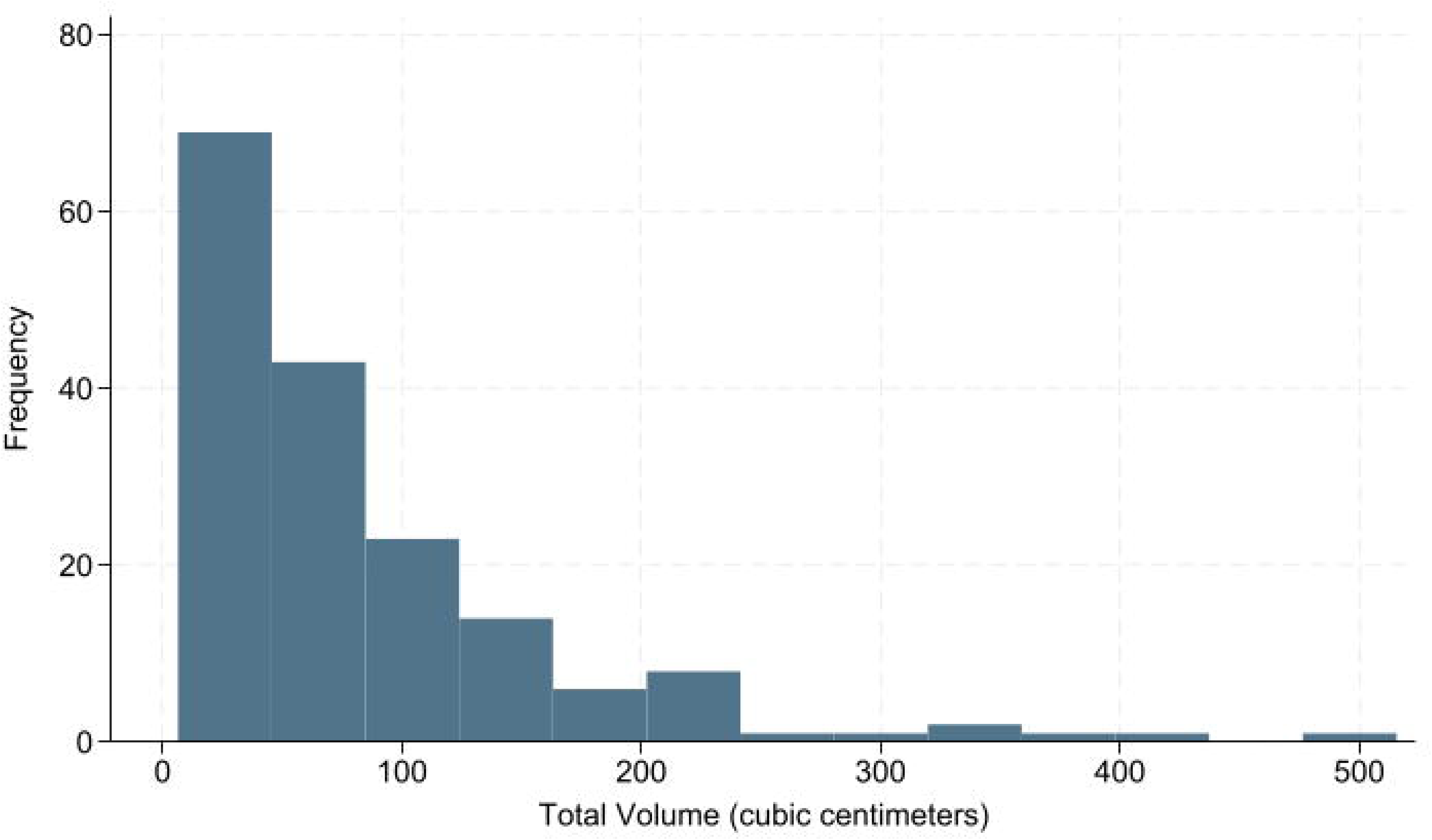
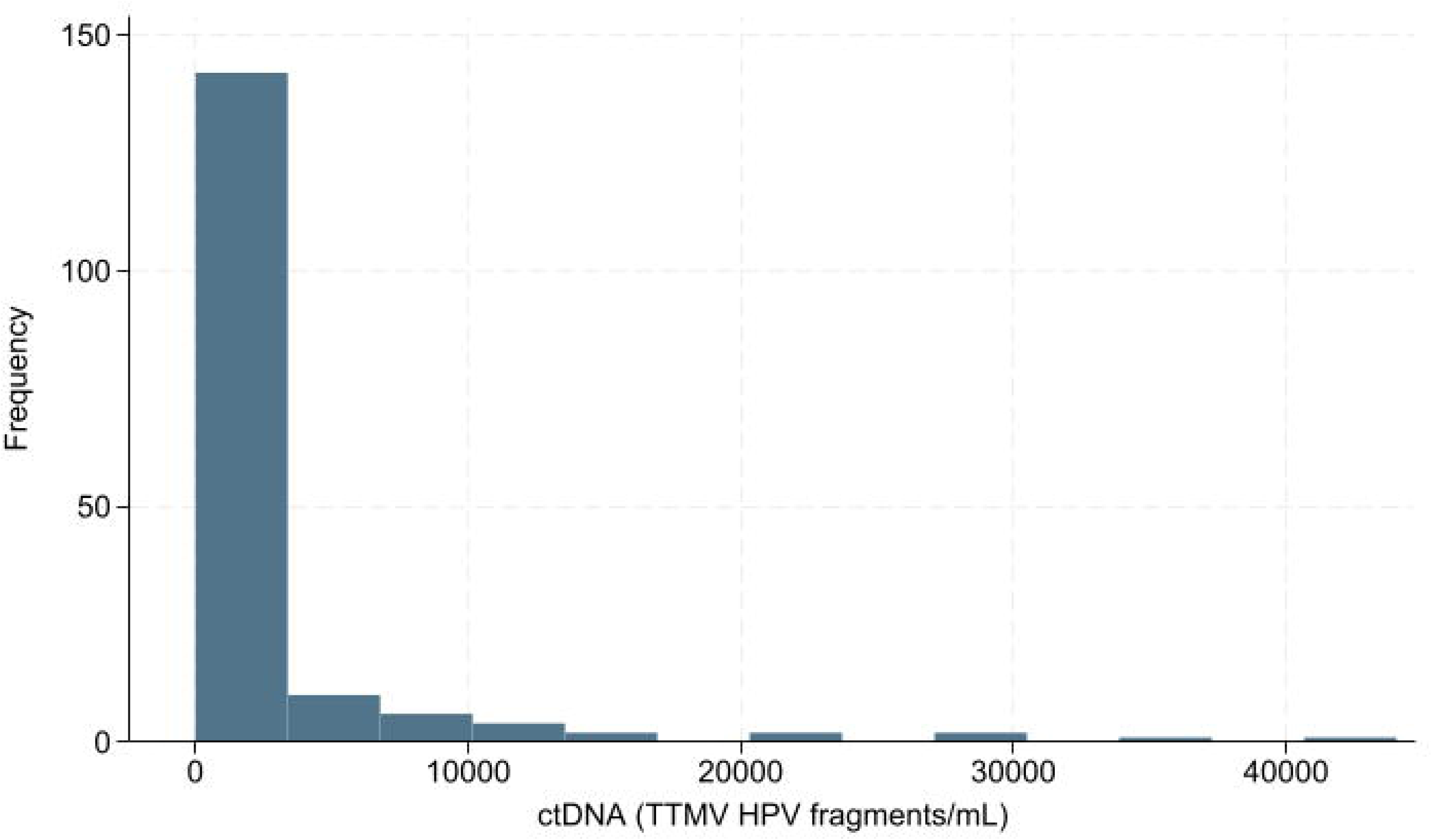
Histograms of Automated Tumor Volume, Automated Nodal Volume, Automated Total Volume (cubic centimeters), and circulating tumor DNA (tumor-tissue modified HPV fragments/mL)

**Table 1.**
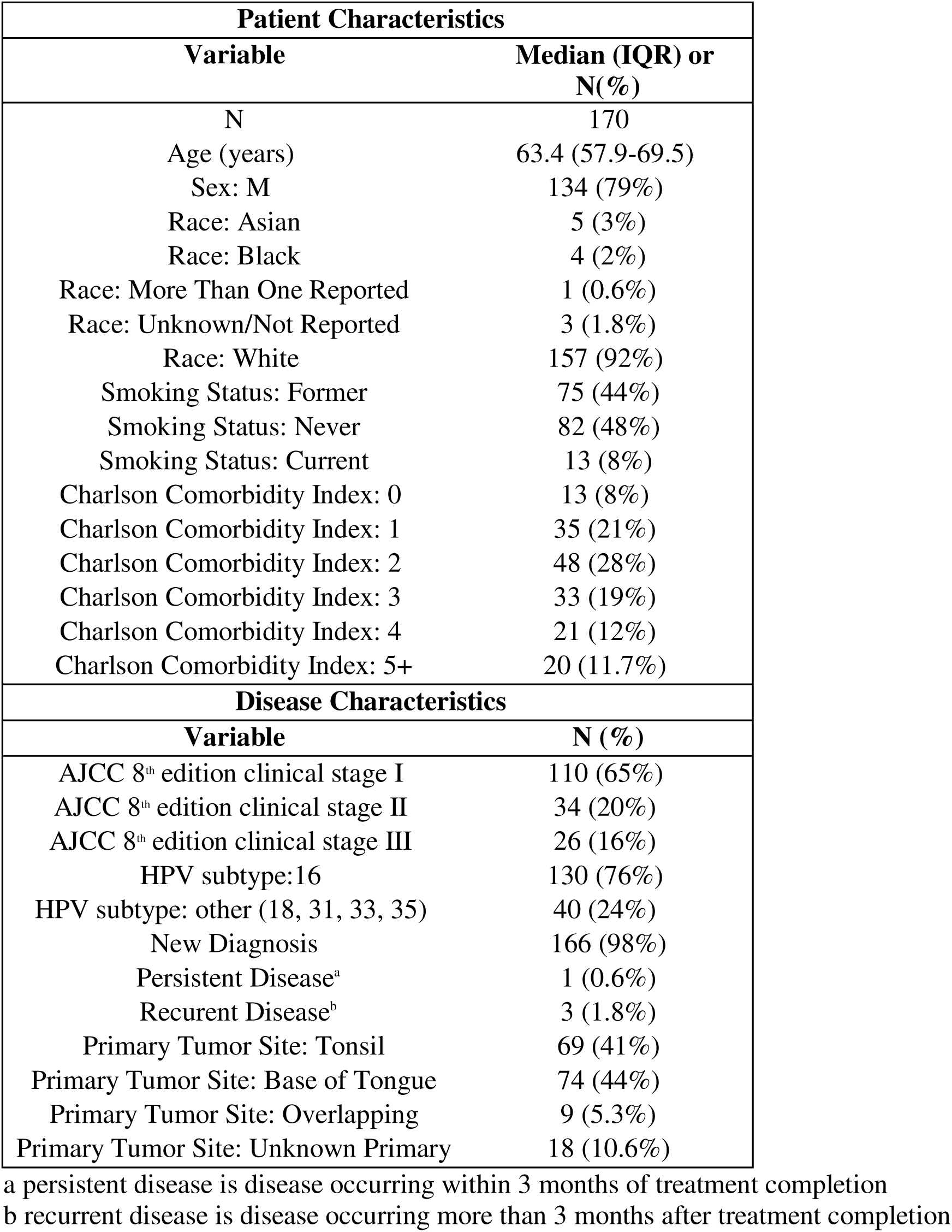
Patient and Disease Characteristics.

**Table 2.** Automated Volumes and CtDNA.

| Variable | Mean (SD) |
| --- | --- |
| ctDNA (TTMV<sup>a</sup> HPV Fragments/mL) | 2499.27 (6189.78) |
| Automated tumor volume (cc) | 42.56 (57.99) |
| Automated nodal volume (cc) | 41.35 (45.04) |
| Automated total volume (cc) | 83.91 (80.56) |
| Automated cystic<sup>b</sup> nodal volume (cc) | 3.37 (7.46) |
| Automated cystic nodal ratio<sup>c</sup> | 0.10 (0.12) |

a TTMV: tumor tissue modified viral DNA
b Cystic or internally necrotic matter was measured internal lymph node hypodensity within the range from -100 to +20 Hounsfield Units
c Cystic nodal ratio was calculated as the ratio of volume of internal lymph node hypodensity/total nodal volume

Two-way scatter plots were generated to illustrate the association between ctDNA and automated primary tumor, nodal and total volumes (**Figures 3a-3c**). Spearman correlation testing demonstrated a low magnitude but statistically significant correlation between AI-auto-segmented primary tumor volume and ctDNA (ρ=0.28, p<0.001), and between ctDNA and AJCC 8^th^ edition clinical tumor stage (ρ=0.25, p<0.001). Auto-segmented nodal volume demonstrated a moderate, statistically significant correlation with ctDNA (ρ=0.53, p<0.001) which was greater in magnitude than the correlation with clinical nodal staging (ρ=0.45, p<0.001). There was a significant correlation between ctDNA and total auto-segmented volume (ρ=0.56, p<0.001) which was greater in magnitude than the significant association between ctDNA and overall clinical stage (ρ=0.35, p<0.001). There was a modest but statistically significant correlation between ctDNA and cystic nodal volume (ρ=0.28, p<0.001), and a low but inverse correlation between ctDNA and cystic ratio (ρ=-0.20, p=0.009) (**Figure 4**).

**Figure 3a-3c.**
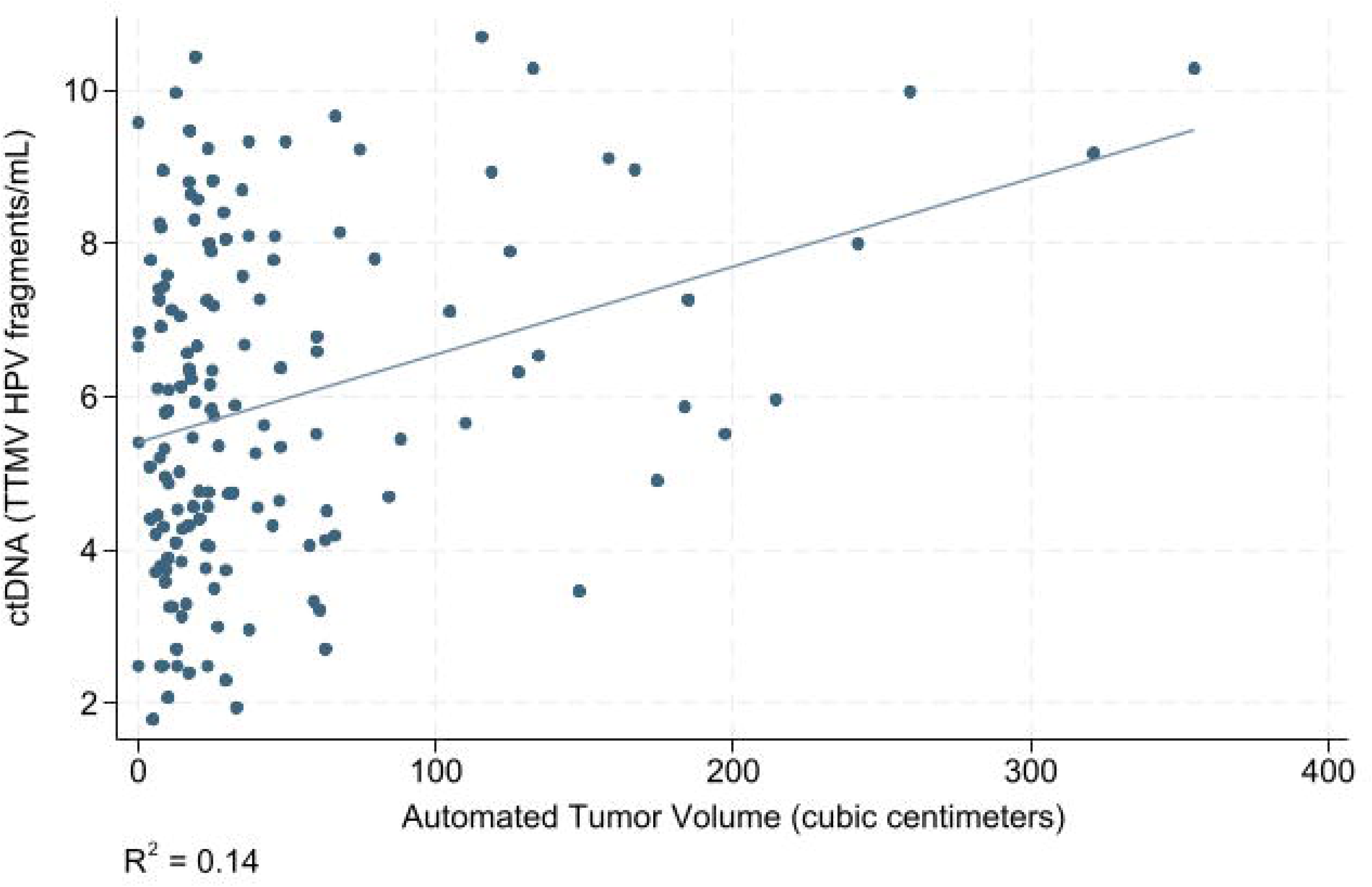
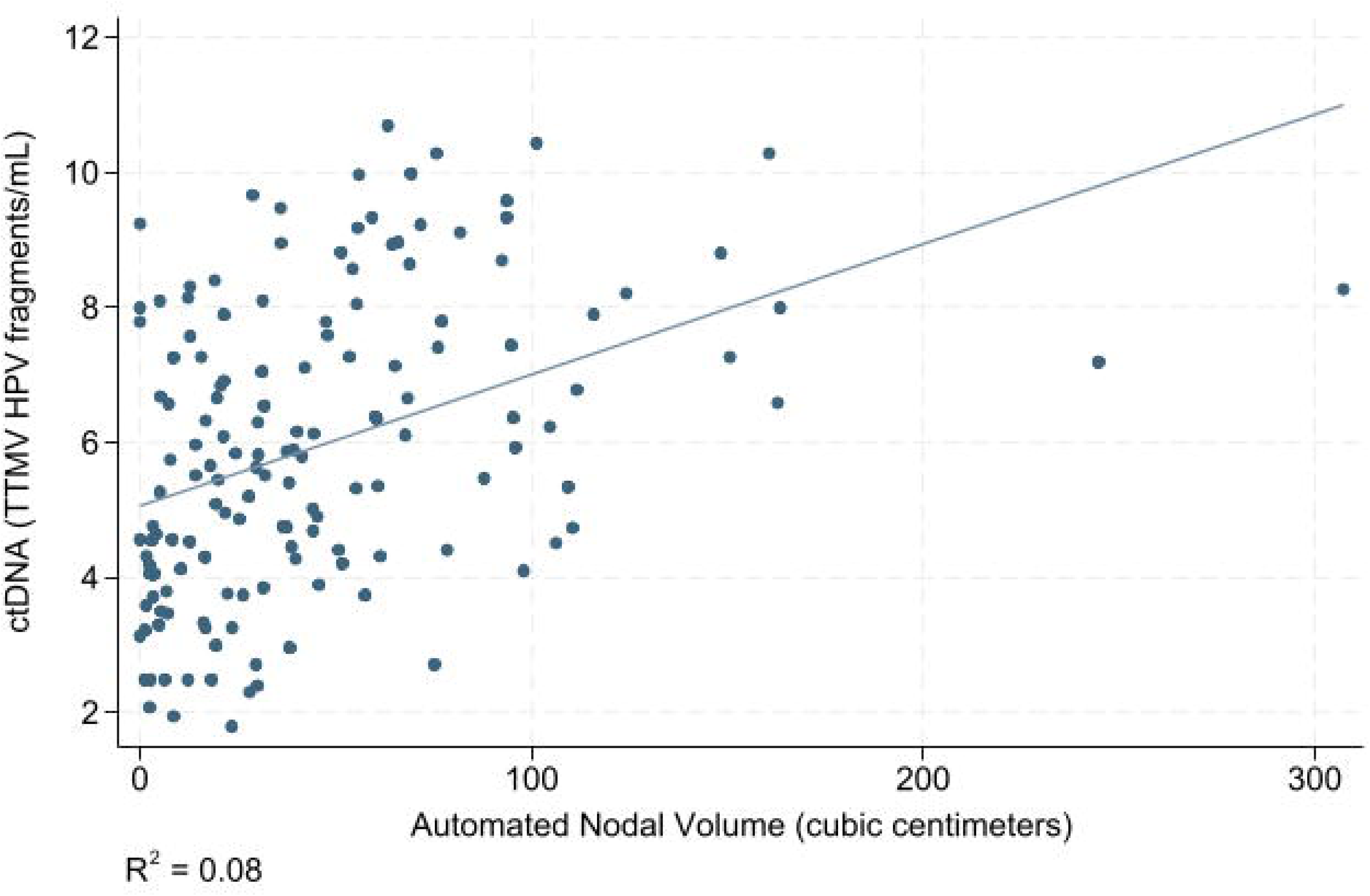
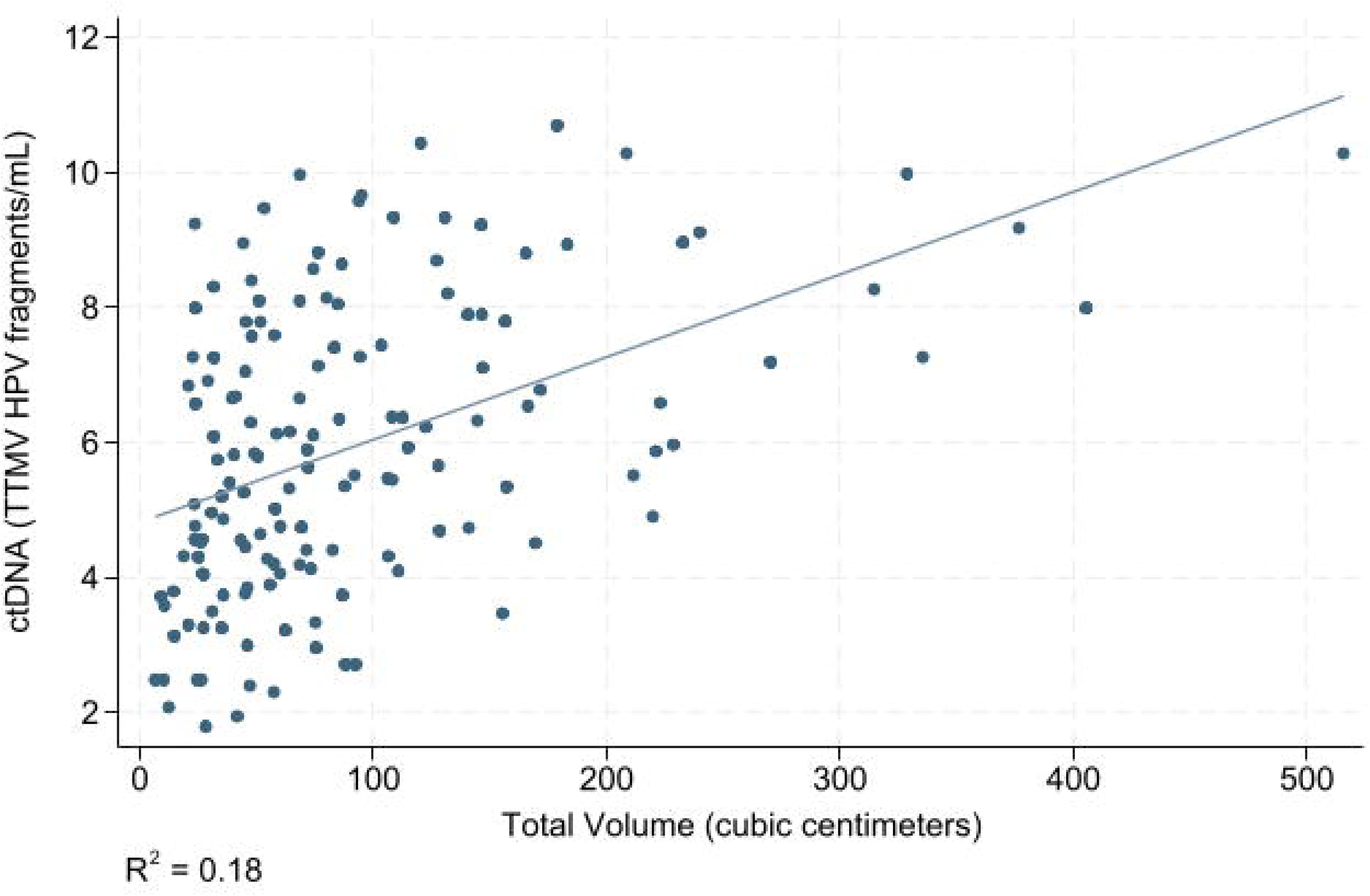
Scatter Plots of Automated Tumor Volume, Automated Node Volume, Automated Total Volume (cubic centimeters), against log (circulating tumor DNA)^a^ (tumor-tissue modified HPV fragments/mL) a ctDNA has been log-transformed

**Figure 4.**
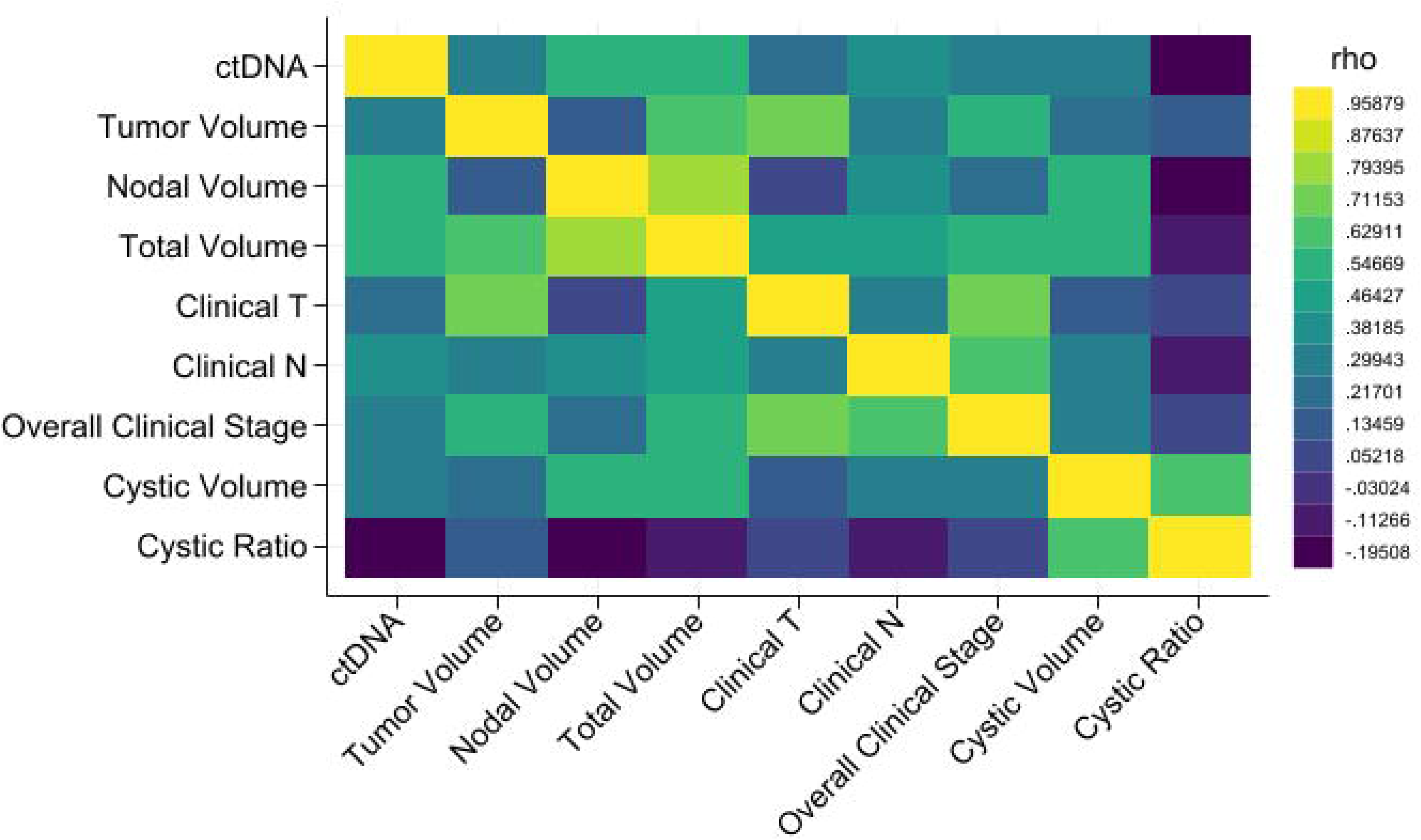
Spearman Correlation Testing Heat Map.

In the univariable linear regression, ctDNA was associated with auto-segmented primary tumor volume (coeff=39.43, p<0.001), and with auto-segmented nodal volume (coeff =39.54, p<0.001). Additionally, ctDNA was associated with AJCC 8^th^ edition clinical T stage (coeff=1031.09, p=0.009) and AJCC 8^th^ edition clinical N stage (coeff=1840, p=0.018). Automated volume measurement of cystic or necrotic appearing material, and the ratio of cystic or necrotic fluid to node volume were not significantly associated with ctDNA in the univariable regression (p=0.11, p=0.16, respectively). When calculated using an alternative HU threshold of −20 to +20 HU, the results were similar (**Supplementary Table 1**). HPV-16 vs other subtypes (coeff=3072.40, p=0.006) was also significantly associated with ctDNA. Smoking status as former smoker (coeff=-6126.85, p=0.001) or never smoker (coeff =-5659.70, p=0.002) were negatively associated with ctDNA when compared to current smoker, as was CCI (−596.60, p=0.038). Age in years and sex were not significantly associated with ctDNA (p>0.05) (**Table 3a).**

**Table 3a.** Single Variable Regression.

| Variable | Coefficient | Std. Error | p value |
| --- | --- | --- | --- |
| Automated tumor volume | 39.43 | 7.65 | <0.001 |
| Automated nodal volume | 39.54 | 10.15 | <0.001 |
| Cystic node volume | 0.31 | 0.19 | 0.11 |
| Cystic node ratio | -5912.99 | 4137.31 | 0.16 |
| AJCC 8<sup>th</sup> edition clinical T stage | 1031.09 | 392.26 | 0.009 |
| AJCC 8<sup>th</sup> edition clinical N stage | 1840 | 770.99 | 0.018 |
| Age (years) | -79.32 | 47.92 | 0.10 |
| Male sex | 1443.57 | 1160.09 | 0.22 |
| Female sex | Reference | Reference | Reference |
| Smoking status: Former | -6126.85 | 1808.39 | 0.001 |
| Smoking status: Never | -5659.70 | 1796.95 | 0.002 |
| Smoking status: Current | Reference | Reference | Reference |
| CCI<sup>a</sup> | -596.60 | 285.66 | 0.038 |
| HPV subtype = 16 | 3072.40 | 1097.19 | 0.006 |
| HPV subtype = other<sup>b</sup> | Reference | Reference | Reference |

a CCI: Charlson Comorbidity Index, measured ordinally
b included HPV subtypes 18, 31, 33, 35

In the multivariable analysis, while adjusting for smoking status, HPV subtype and CCI, automated tumor and nodal volumes remained significantly associated with ctDNA (coeff=34.79, p=0.001; coeff=24.68, p=0.022 respectively). Meanwhile, in the multivariable analysis, AJCC 8^th^ edition clinical T stage and clinical N stage were no longer statistically significantly associated with ctDNA when controlling for these clinical factors (p=0.37 and p=0.29, respectively) (**Table 3b**).

**Table 3b.** Multivariable Regression Including Automated Tumor and Nodal Volumes.

| **Variable** | **Coefficient** | **Std Error** | **p value** |
| --- | --- | --- | --- |
| Automated tumor volume | 34.79 | 10.40 | 0.001 |
| Automated nodal volume | 24.68 | 10.67 | 0.022 |
| Smoking status: Former | -3657.39 | 1745.85 | 0.038 |
| Smoking status: Never | -2738.99 | 1736.14 | 0.12 |
| Smoking status: Current | Reference | Reference | Reference |
| HPV type: 16 | 1788.19 | 1035.73 | 0.086 |
| HPV type: other | Reference | Reference | Reference |
| CCI | -433.46 | 262.65 | 0.10 |
| AJCC 8<sup>th</sup> edition clinical T stage | -439.28 | 490.96 | 0.37 |
| AJCC 8<sup>th</sup> edition clinical N stage | 238.19 | 809.70 | 0.29 |

Akaike's Information Criterion: 3420.96
Bayesian Information Criterion: 3449.18
Adjusted R<sup>2</sup>: 0.20

Two separate multivariable models were generated, one of which included automated volumes and one which included only clinical tumor and nodal stage (**Figures 5a and 5b**). In the assessment of model fit, The model that included automated volumes demonstrated improved fit, with an Akaike Information Criterion (AIC) of 3420.96 a Bayesian Information Criterion (BIC) of 3449.18, and an adjusted R^2^ of 0.20, compared to the model with only clinical stage (AIC of 3435.88, BIC of 3457.83, and adjusted R^2^ of 0.12) **(Tables 3b and 3c).**

**Figure 5a.**
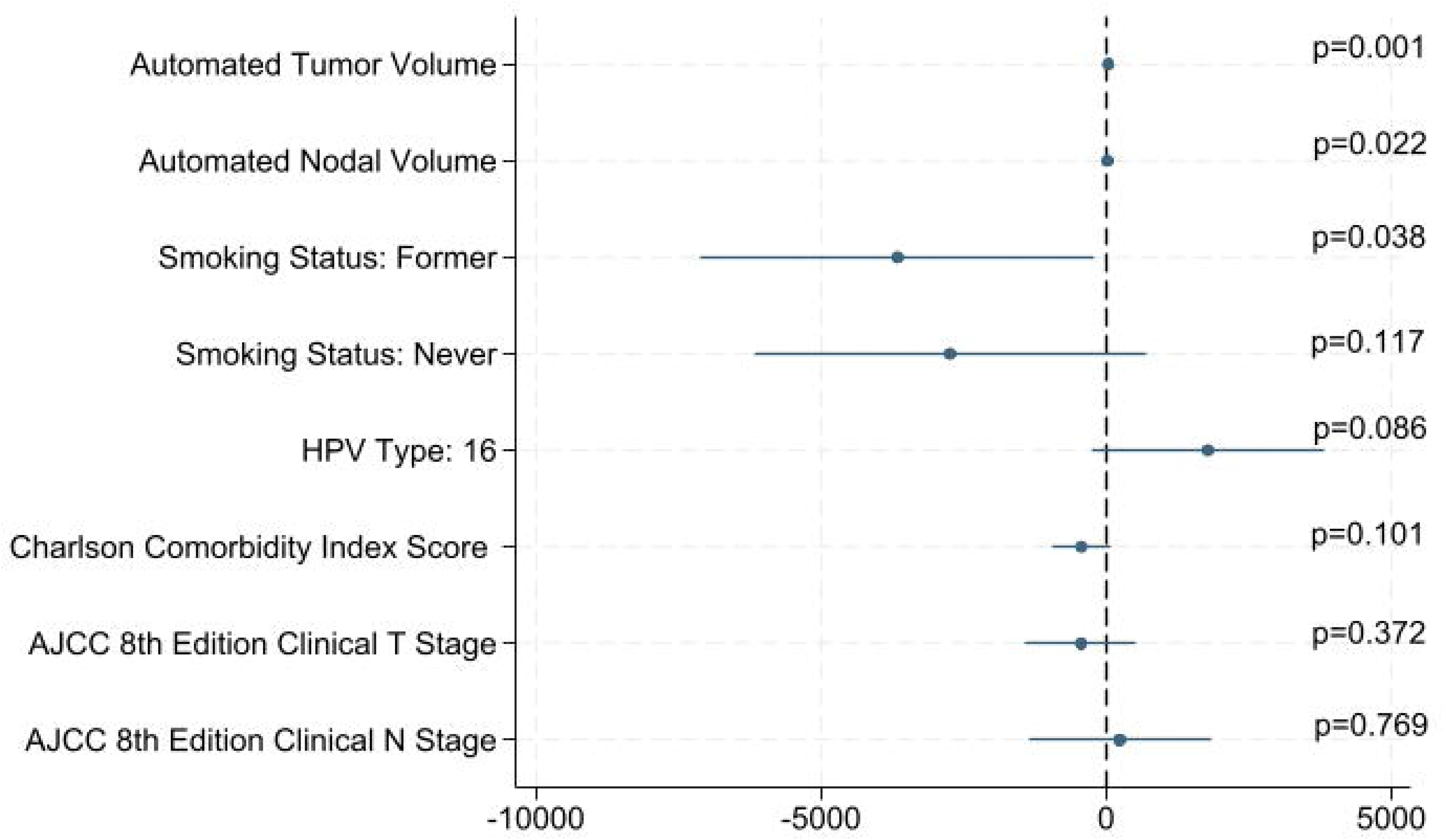
Forest Plot, Multivariable Regression Including Automated Tumor and Nodal Volumes.

**Figure 5b.**
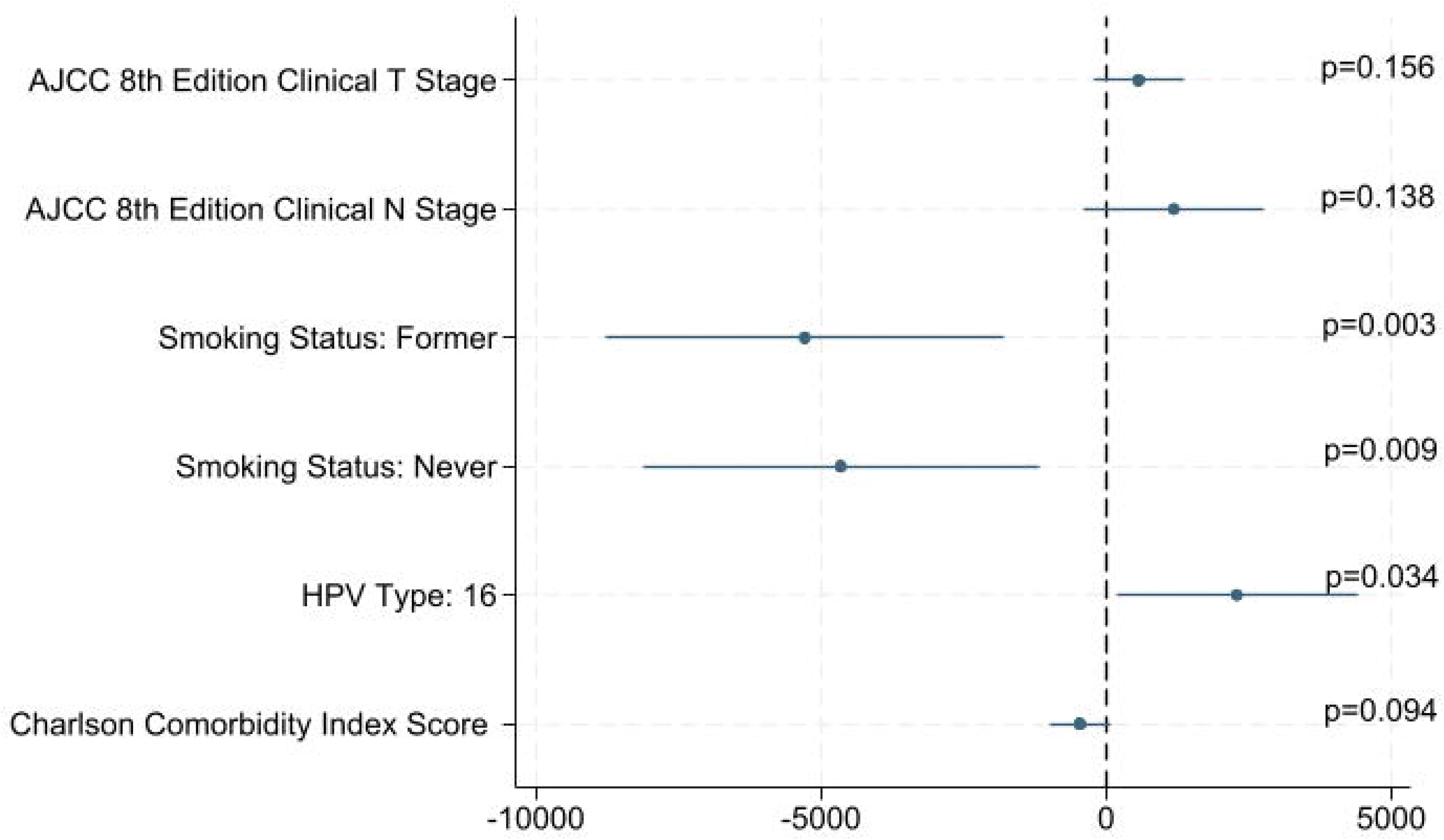
Forest Plot, Multivariable Regression Omitting Automated Tumor and Nodal Volumes.

**Table 3c.**
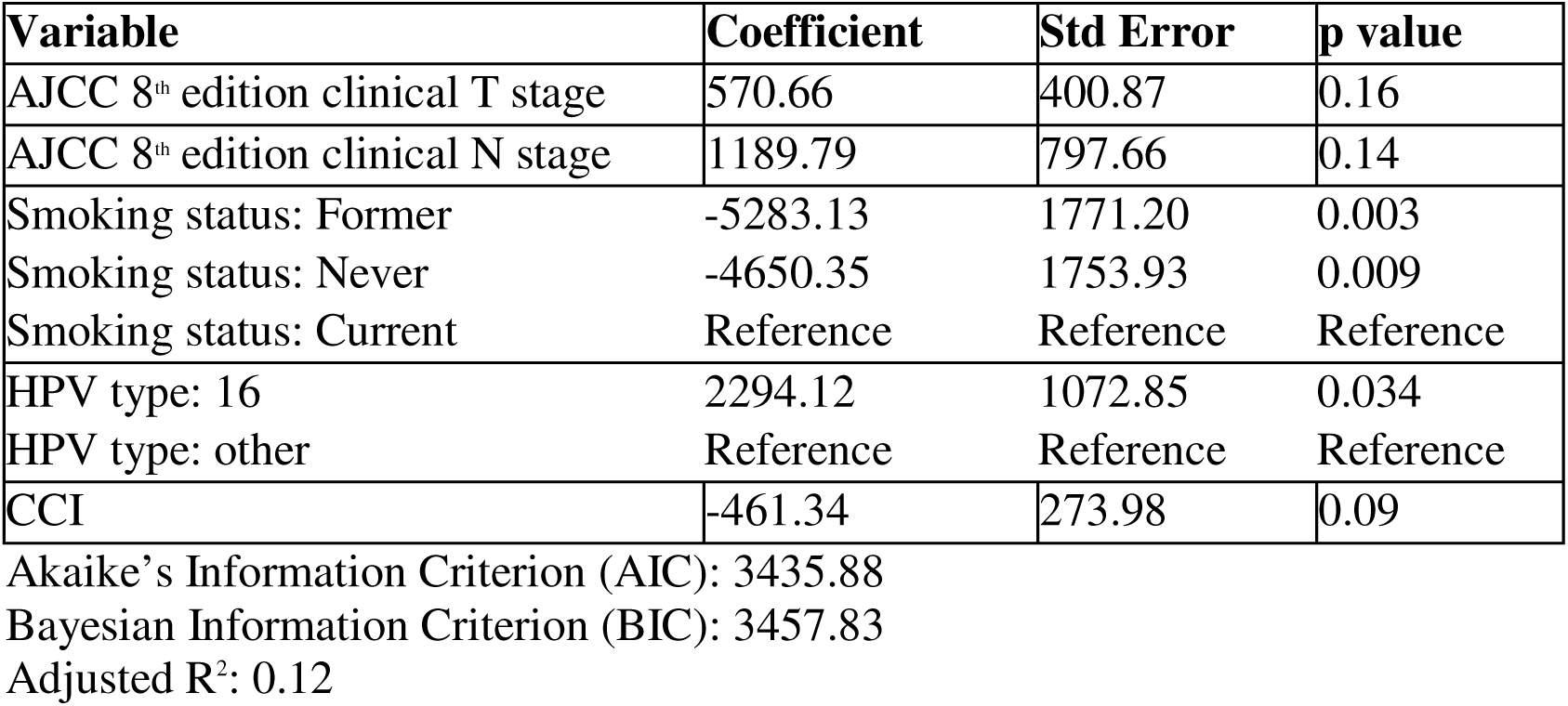
Multivariable Regression Omitting Automated Tumor and Nodal Volumes.

### Subgroup Analyses

Two exploratory subgroup analyses were conducted. For each subgroup, a Bonferroni correction was applied to set the significance threshold at 0.005 for 10 primary comparisons in the univariable analysis, and at 0.0083 for 6 predictor variables in the multivariable analysis.

#### ctDNA and HPV subtype

Forty patients (24%) included in the study tested positive for non-HPV-16 subtypes (18, 31, 33 and 35). We conducted an exploratory subgroup analysis of the association between ctDNA and automated tumor burden within this group.

In the univariable analysis, automated tumor volume was no longer significantly associated with ctDNA (coeff=1.79, p=0.19), while automated nodal volume remained significantly associated with ctDNA (coeff=8.26, p<0.001) (**Supplementary Figures 1a-1c**). Cystic or necrotic node volume and cystic node ratio were not significantly associated with ctDNA (p=0.34 and p=0.17, respectively). AJCC 8^th^ edition clinical T stage, clinical N stage, age, sex, smoking status, and CCI were not significantly associated with ctDNA in this subgroup.

In the multivariable analysis, automated tumor volume was no longer significantly associated with ctDNA (p=0.57), but automated nodal volume remained significantly associated (coeff = 8.89, p<0.001). AJCC 8^th^ edition clinical T or N stage, smoking status, and CCI were no longer significantly associated with ctDNA (**Supplementary Table 2a**).

#### High-volume nodal disease

An exploratory subgroup analysis was also conducted for patients with automated nodal volumes measuring greater than the 80^th^ percentile, or 67.8 cubic centimeters.

In the univariable analysis, automated tumor volume was significantly associated with ctDNA (coeff=53.52, p=0.003), while automated nodal volume was not (coeff=-12.61, p=0.70). Cystic node volume and cystic ratio were not significantly associated (p=0.95 and p=0.67, respectively). AJCC 8^th^ edition clinical T stage and clinical N stage were not significantly associated with ctDNA in this subgroup (p=0.21 and p=0.35, respectively). Smoking status as former smoker (coeff =-16713.89, p=0.001) or never smoker (coeff=-14636.02, p=0.003) remained significantly associated with ctDNA. None of the remaining clinical factors tested were significantly associated with ctDNA.

In the multivariable model, ctDNA was no longer significantly associated with nodal volume (coeff=-8.43, p=0.76) and remained significantly associated with automated tumor volume (coeff=56.86, p=0.014). Smoking status of former smoker (coeff=-12677.35, p=0.01) or never smoker (coeff=-11051.18, p=0.02) and CCI (coeff=-1773, p=0.05) also remained significantly associated with ctDNA. HPV subtype and AJCC 8^th^ edition clinical stage were no longer associated with ctDNA (**Supplementary Table 2b**).

## Discussion

This study adds to the existing literature as the first use of Artificial Intelligence (AI) driven volumetric assessment of tumor burden to complement ctDNA. Circulating HPV DNA has demonstrated promise in informing follow up of patients with HPV-OPSCC^10^ and a growing number of studies are evaluating the relationship between ctDNA and disease burden and whether ctDNA assessment can be used to guide patient management, particularly to risk-stratify patients for de-escalated therapy.

Circulating HPV tumor DNA has been previously associated with tumor burden via clinical staging or radiologist/radiation oncologist assessment of imaging. For instance, Agarwal et al.^16^ demonstrated a strong concordance between ctDNA values and PET-CT images for patients under surveillance for HPV. Similarly, Cao et al.^17^ compared the predictive value between ctDNA and imaging biomarkers in both MRI and FDG-PET images collected pre-treatment and during chemoradiation. The authors identified multiple imaging parameters, including metabolic tumor volume on FDG-PET, which demonstrate a strong correlation with ctDNA. In addition to tumor and nodal staging, other imaging biomarkers, including tumor metabolism and hypoxia, have been associated with ctDNA and disease response.^17^ Moreover, volumetric analysis of imaging such as PET has also been associated with locoregional failure free survival in HPV-OPSCC.^18^ These studies set a precedent for further exploration of the association between imaging biomarkers and HPV DNA liquid biopsy, but with limitations including small sample size and manual imaging interpretation.

Alternatively, Artificial Intelligence (AI)-driven imaging analysis offers a burgeoning area of research in radiomics and a method for quickly and practically delineating tumor and nodal volumes without manual input. In HPV-OPSCC, AI deep-learning algorithms have proven useful in detecting baseline disease characteristics such as gross tumor volume^19^ and extra-nodal extension^20^ as well as in auto-segmentation to assist with radiation treatment planning.^21–23^ The association between AI-generated volumetrics and ctDNA in HPV-associated oropharynx cancers remains to be explored.

Auto-segmentation has an expanding role in head and neck cancers. Prior investigations have applied deep learning algorithms to auto-segment normal tissue structures, gross tumor volumes (GTV),^24^ clinical tumor volumes (CTV)^25^ and lymph nodes^26^ for radiotherapy planning. Our study adds to the clinical applications of deep-learning auto-segmentation by demonstrating associations between auto-segmented tumor volumes and ctDNA. Automated imaging analysis has enormous practical benefit because it allows for real-time assessment of disease and addresses both the time limitations and potentially the cost constraints as a complementary measure of ctDNA. In the future, auto-segmentation may be integrated into the clinical setting, but would require implementation via user-friendly and widely accessible clinical tools. This could become increasingly important as ctDNA is investigated for pre-treatment risk-stratification, but also in the assessment of disease response and surveillance for recurrence.^7^ In other studies, imaging analysis has correlated with ctDNA throughout the disease course^17^; this is yet to be determined for AI-generated volumetrics.

In our study, we found the strongest correlation between the automated volume of nodal disease and ctDNA levels, consistent with other prior studies that have identified nodal disease burden as a predictor of ctDNA levels.^5^ This relationship warrants further study, as the biologic mechanisms by which nodal disease preferentially gives rise to ctDNA versus primary tumor volume is unknown. Additionally, we identified a potential threshold, in the upper quintile of nodal volume, where this relationship no longer holds. We are currently conducting translational studies to evaluate this further.^6^

In this study, we hypothesize that cystic, or internally necrotic lymph node regions may correlate with cellular turnover and DNA spread; however, we find that there is no association between cystic nodal volumes when measured through two separate Hounsfield Unit thresholds. There is a subtle inverse correlation between ctDNA and cystic ratio, suggesting that a greater cystic component of lymph nodes may serve to sequester DNA and prevent its spread.

Finally, there are additional subgroups for whom ctDNA may not always correlate with tumor burden. In our study, patients belonging to the non-HPV-16 subgroup (with genotypes 18, 31, 33, and 35), showed no significant association between automated tumor volumes and ctDNA, and still showed a statistically significant, but lower effect size, association between nodal volumes and ctDNA. While HPV-16 remains the most common subtype associated with HPV-OPSCC in the United States, ctDNA assays may now detect a variety of genotypes and it is important to explore how the relationship between tumor burden and ctDNA may differ in those with non-HPV-16 driven disease, and whether this has any additional prognostic value.

Given the demonstrated associations between ctDNA and radiographic features, there is growing interest in integrating imaging and circulating biomarkers to predict cancer outcomes.^27–31^

## Limitations

There are several limitations to this study. First, this is a secondary analysis of a single-institution prospective cohort. Further validation of AI-generated volumetrics would require prospective application to an external or multi-institutional cohort of patients. Second, we applied the auto-segmentation algorithm to a combination of pre-treatment scans, including diagnostic CT, diagnostic PET-CT, and pre-radiation CT simulation scans. This may not account for differences between imaging type and quality; a more accurate assessment of the relationship between automated volumetrics and ctDNA may require application to a single type of imaging. Finally, our study is limited to the pre-treatment setting, and further investigation is needed to explore the associations between AI-generated volumetrics and ctDNA during treatment and in recurrent or metastatic disease. Notably, the patients analyzed in this study were enrolled in a prospective protocol investigating the use of ctDNA to drive post-treatment surveillance in HPV-OPSCC.

## Conclusion

In this study, we demonstrate the association between AI-generated volumetrics on pre-treatment imaging and ctDNA. The association is stronger than AJCC 8^th^ edition clinical staging in single and multivariable analysis. Inclusion of AI-generated volumetrics improved the predictive capacity of multivariable models. Automated imaging analysis may provide a practical, cost-effective way to measure tumor burden in real time and provide a new avenue for risk stratification for de-escalated therapy in HPV-OPSCC. Further investigation into AI-generated volumetrics and liquid biomarkers in HPV-OPSCC is warranted.

## Data Availability

All data produced in the present study are available upon reasonable request to the authors.

## Figure titles, in order of appearance

**Supplementary Table 1.**
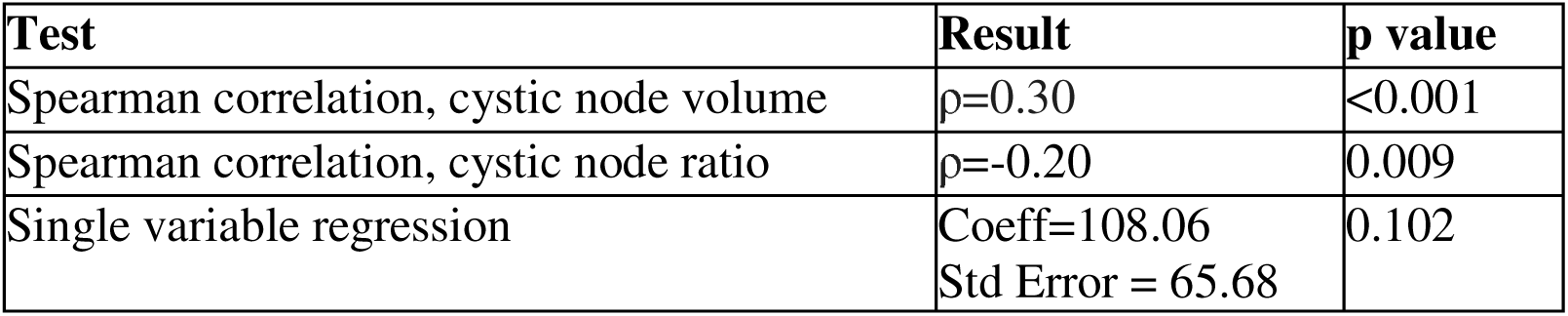
Cystic Node Volume Method 2 [−20] to [+20] HU.

**Supplementary Table 2a.** Subgroup Analysis for non-HPV-16 subtypes.

| Single Variable Analysis |  |  |  |
| --- | --- | --- | --- |
| Variable | Coefficient | Std Error | p value |
| Automated tumor volume | 1.79 | 1.35 | 0.19 |
| Automated nodal volume | 8.26 | 1.37 | <0.001 |
| Cystic node volume | 0.04 | 0.04 | 0.34 |
| Cystic node ratio | -450.88 | 320.39 | 0.17 |
| AJCC 8<sup>th</sup> edition clinical T stage | 37.44 | 49.26 | 0.45 |
| AJCC 8<sup>th</sup> edition clinical N stage | 177.99 | 85.11 | 0.043 |
| Age (years) | 3.89 | 6.01 | 0.52 |
| Male sex | -76.70 | 110.78 | 0.49 |
| Female sex | Reference | Reference | Reference |
| Smoking status: Former | -140.76 | 248.40 | 0.57 |
| Smoking status: Never | -258.83 | 245.90 | 0.30 |
| Smoking status: Current | Reference | Reference | Reference |
| CCI | -7.46 | 34.67 | 0.83 |
| Multivariable Analysis |  |  |  |
| Variable | Coefficient | Std Error | p value |
| Automated tumor volume | 0.92 | 1.61 | 0.57 |
| Automated nodal volume | 8.89 | 1.61 | <0.001 |
| Smoking status: Former | -178.32 | 208.34 | 0.40 |
| Smoking status: Never | -228.79 | 187.51 | 0.23 |
| Smoking status: Current | Reference | Reference | Reference |
| CCI | 20.64 | 28.63 | 0.48 |
| AJCC 8<sup>th</sup> edition clinical T stage | 25.80 | 49.60 | 0.61 |
| AJCC 8<sup>th</sup> edition clinical N stage | -49.42 | 91.96 | 0.60 |

**Supplementary Table 2b.** Subgroup Analysis for High Volume Nodal Disease.

| **Single Variable Regression** |  |  |  |
| --- | --- | --- | --- |
| **Variable** | **Coefficient** | **Std Error** | **p value** |
| Automated tumor volume | 53.52 | 16.72 | 0.003 |
| Automated nodal volume | -12.61 | 31.85 | 0.70 |
| Cystic node volume | -0.02 | 0.33 | 0.95 |
| Cystic node ratio | 10755.94 | 24627.12 | 0.67 |
| AJCC 8<sup>th</sup> edition clinical T stage | 1444.90 | 1122.22 | 0.21 |
| AJCC 8<sup>th</sup> edition clinical N stage | -2318.20 | 2428.96 | 0.35 |
| Age (years) | -77.90 | 146.43 | 0.60 |
| Male sex | 5441.13 | 4091.01 | 0.19 |
| Female sex | Reference | Reference | Reference |
| Smoking status: Former | -16713.89 | 4325.60 | 0.001 |
| Smoking status: Never | -14636.02 | 4450.56 | 0.003 |
| Smoking status: Current | Reference | Reference | Reference |
| CCI | -1663.81 | 1064.44 | 0.13 |
| **Multivariable Analysis** |  |  |  |
| Automated tumor volume | 56.86 | 21.64 | 0.01 |
| Automated nodal volume | -8.43 | 26.87 | 0.76 |
| Smoking status: Former | -12677.35 | 4600.46 | 0.01 |
| Smoking status: Never | -11051.18 | 4581.42 | 0.02 |
| Smoking status: Current | Reference | Reference | Reference |
| CCI | -1773 | 852.86 | 0.05 |
| AJCC 8<sup>th</sup> edition clinical T stage | -1764.13 | 1329.95 | 0.20 |
| AJCC 8<sup>th</sup> edition clinical N stage | -391.22 | 2236.82 | 0.86 |

**Supplementary Figure 1.**
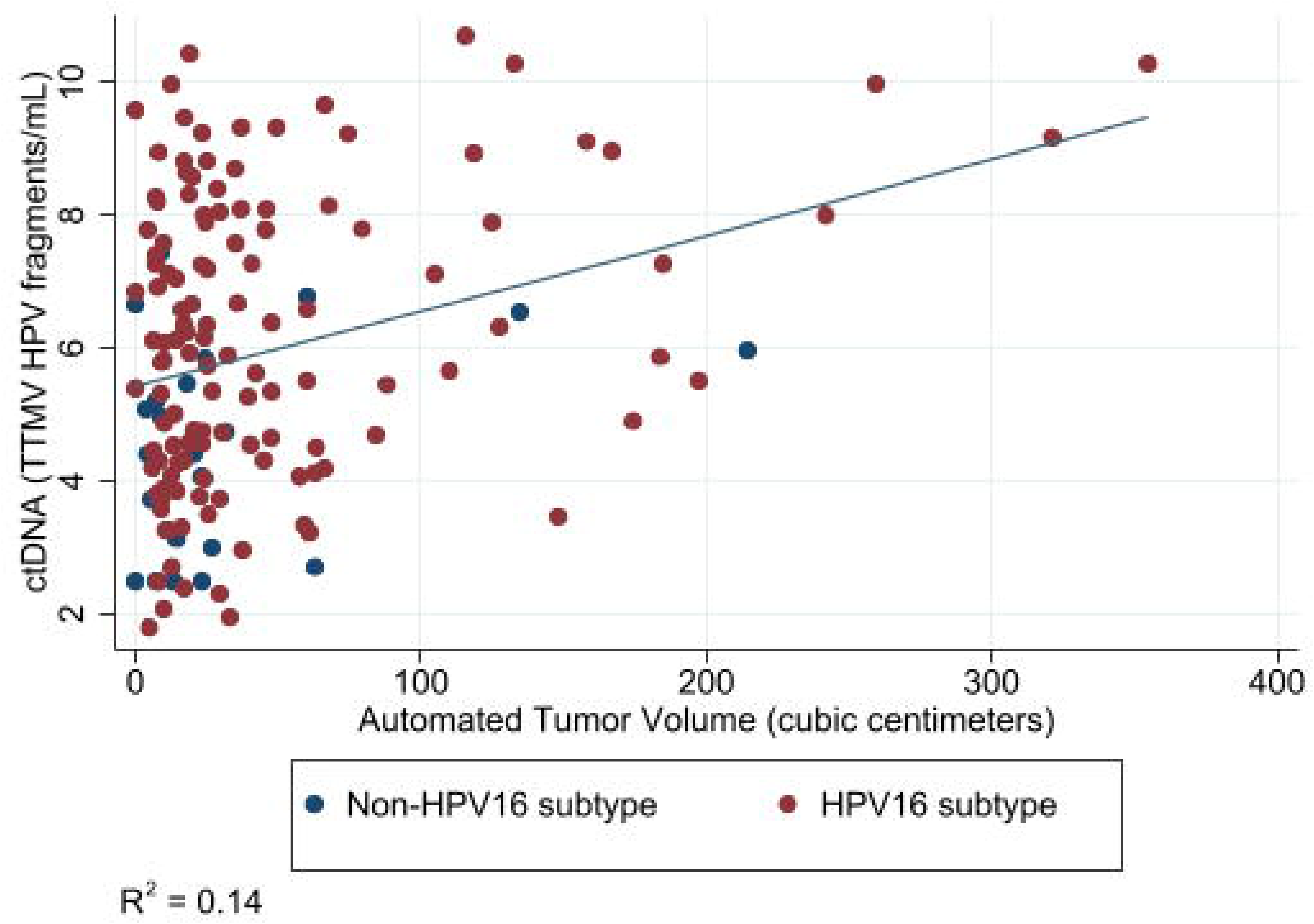
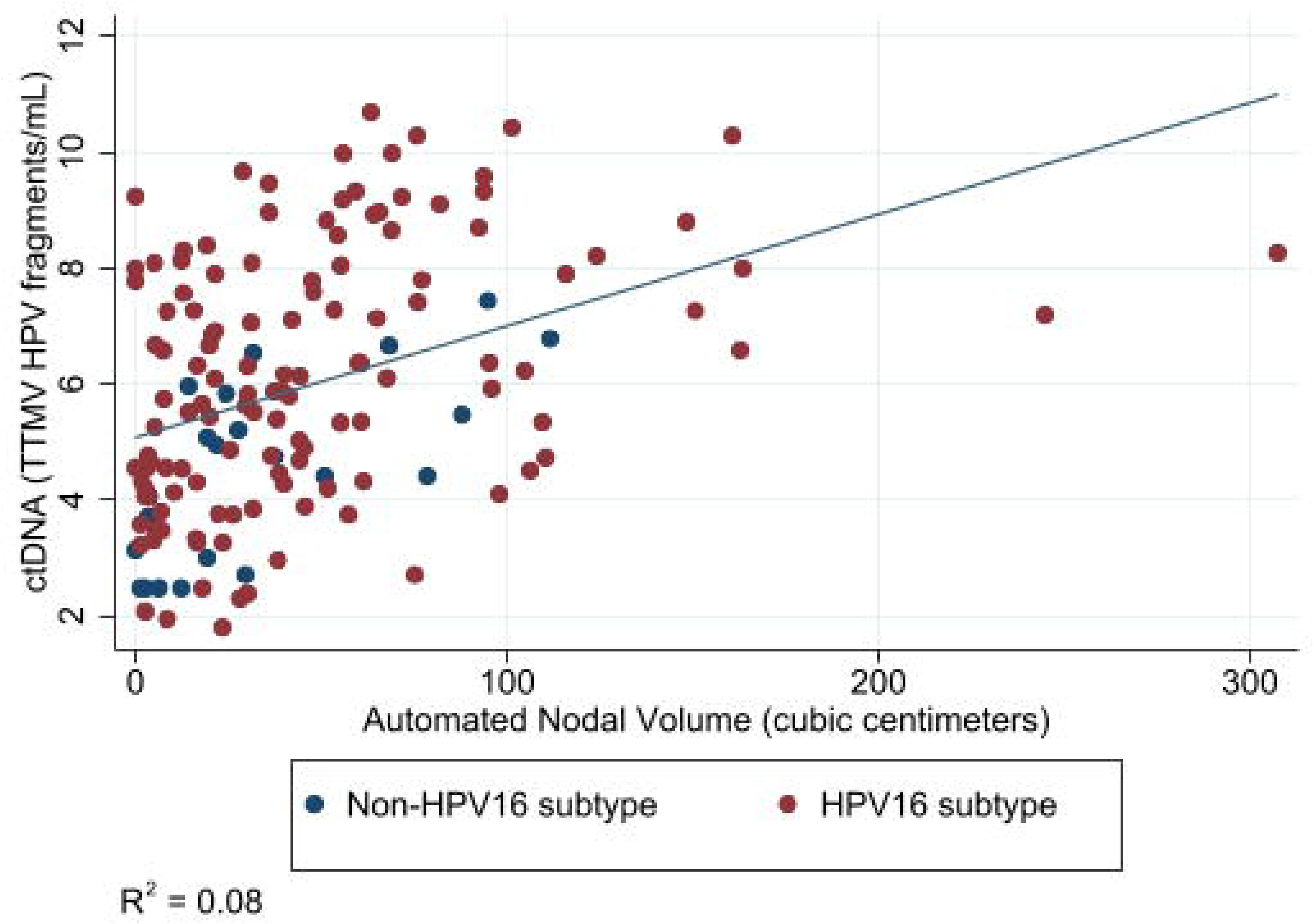
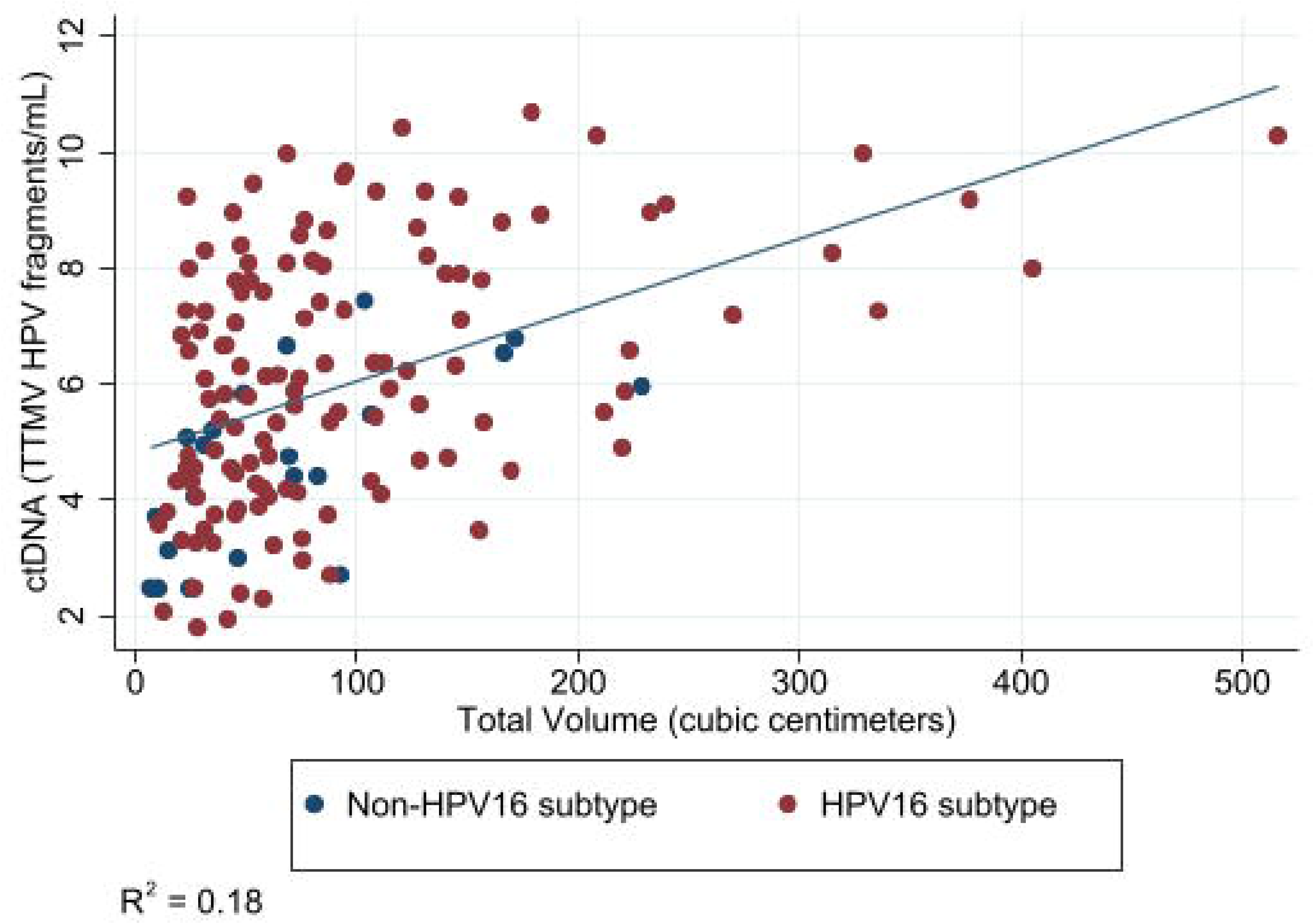
Scatter Plots of Automated Tumor Volume, Automated Nodal Volume, Automated Total Volume (cubic centimeters), against log (circulating tumor DNA)^a^ (tumor-tissue modified HPV fragments/mL) by HPV genotype (16 vs other) a ctDNA has been log-transformed

## References

1. Centers for Disease Control: Cancers Linked with HPV Each Year. https://www.cdc.gov/cancer/hpv/cases.html.

2. Ferrandino RM, Chen S, Kappauf C, Barlow J, Gold BS, Berger MH, Westra WH, Teng MS, Khan MN, Posner MR, Misiukiewicz KJ, Bakst RL, Sindhu KK, Genden EM, Chai RL, Roof SA. Performance of Liquid Biopsy for Diagnosis and Surveillance of Human Papillomavirus–Associated Oropharyngeal Cancer. JAMA Otolaryngology–Head & Neck Surgery. 2023;149(11):971. doi:10.1001/jamaoto.2023.1937

3. Aye L, Bryan ME, Das D, Hirayama S, Al-Inaya Y, Mendel J, Naegele S, Fisch AS, Faquin WC, Sadow P, Chan A, Lawrence MS, Mirabello L, Iafrate J, Waterboer T, Wirth LJ, Richmon J, Faden D. Multi-feature next-generation liquid biopsy for diagnosis and prognosis in HPV-associated head and neck cancer. Journal of Clinical Oncology. 2024;42(16_suppl):6064–6064. doi:10.1200/JCO.2024.42.16_suppl.6064

4. Tewari SR, D’Souza G, Troy T, Wright H, Struijk L, Waterboer T, Fakhry C. Association of Plasma Circulating Tumor HPV DNA With HPV-Related Oropharynx Cancer. JAMA Otolaryngology–Head & Neck Surgery. 2022;148(5):488. doi:10.1001/jamaoto.2022.0159

5. Rettig EM, Wang AA, Tran NA, Carey E, Dey T, Schoenfeld JD, Sehgal K, Guenette JP, Margalit DN, Sethi R, Uppaluri R, Tishler RB, Annino DJ, Goguen LA, Jo VY, Haddad RI, Hanna GJ. Association of Pretreatment Circulating Tumor Tissue–Modified Viral HPV DNA With Clinicopathologic Factors in HPV-Positive Oropharyngeal Cancer. JAMA Otolaryngology–Head & Neck Surgery. 2022;148(12):1120. doi:10.1001/jamaoto.2022.3282

6. Chera BS, Kumar S, Shen C, Amdur R, Dagan R, Green R, Goldman E, Weiss J, Grilley-Olson J, Patel S, Zanation A, Hackman T, Blumberg J, Patel S, Thorp B, Weissler M, Yarbrough W, Sheets N, Mendenhall W, Tan XM, Gupta GP. Plasma Circulating Tumor HPV DNA for the Surveillance of Cancer Recurrence in HPV-Associated Oropharyngeal Cancer. Journal of Clinical Oncology. 2020;38(10):1050–1058. doi:10.1200/JCO.19.02444

7. Hanna GJ, Supplee JG, Kuang Y, Mahmood U, Lau CJ, Haddad RI, Jänne PA, Paweletz CP. Plasma HPV cell-free DNA monitoring in advanced HPV-associated oropharyngeal cancer. Annals of Oncology. 2018;29(9):1980–1986. doi:10.1093/annonc/mdy251

8. Yom SS, Harris J, Caudell JJ, Geiger JL, Waldron J, Gillison M, Subramaniam RM, Yao M, Xiao C, Kovalchuk N, Martino R, Jordan R, Henson C, Echevarria M, Lominska CE, Dorth JA, Stokes WA, Chan J, Gensheimer MF, Le QT. Interim Futility Results of NRG-HN005, A Randomized, Phase II/III Non-Inferiority Trial for Non-Smoking p16+ Oropharyngeal Cancer Patients. International Journal of Radiation Oncology*Biology*Physics. 2024;120(2):S2–S3. doi:10.1016/j.ijrobp.2024.08.014

9. Kowalchuk RO, Kamdem Talom BC, Van Abel KM, Ma DM, Waddle MR, Routman DM. Estimated Cost of Circulating Tumor DNA for Posttreatment Surveillance of Human Papillomavirus–Associated Oropharyngeal Cancer. JAMA Netw Open. 2022;5(1):e2144783. doi:10.1001/jamanetworkopen.2021.44783

10. Rettig EM, Schoenfeld JD, Miller J, Sargent B, Carey E, Margalit DN, Sehgal K, Sethi RKV, Uppaluri R, Tishler RB, Goguen LA, Annino DJ, Sim ES, Jo VY, Wong KS, Guenette JP, Haddad RI, Hanna GJ. A Prospective Trial of Biomarker-Guided Surveillance for HPV-Positive Oropharynx Cancer Using Plasma Tumor Tissue–Modified Viral HPV DNA. Clinical Cancer Research. Published online January 13, 2025:OF1–OF10. doi:10.1158/1078-0432.CCR-24-3053

11. Lowekamp BC, Chen DT, Ibáñez L, Blezek D. The Design of SimpleITK. Front Neuroinform. 2013;7. doi:10.3389/fninf.2013.00045

12. Bradski G. The OpenCV Library. Dr Dobb’s Journal of Software Tools. 2000;25(11).

13. Isensee F, Jaeger PF, Kohl SAA, Petersen J, Maier-Hein KH. nnU-Net: a self-configuring method for deep learning-based biomedical image segmentation. Nat Methods. 2021;18(2):203–211. doi:10.1038/s41592-020-01008-z

14. Jain A, Huang J, Ravipati Y, Cain G, Boyd A, Ye Z, Kann BH. Head and Neck Primary Tumor and Lymph Node Auto-segmentation for PET/CT Scans. In: ; 2023:61–69. doi:10.1007/978-3-031-27420-6_6

15. Lee HN, Han JK, Kim HH, Shin HC, Kim IY, Jou SS. Comparative Study of Lymph Node Metastasis from Squamous Cell Carcinoma and Non-Squamous Cell Carcinoma on Neck CT. Journal of the Korean Society of Radiology. 2015;72(4):271. doi:10.3348/jksr.2015.72.4.271

16. Agarwal A, Bhatt A, Patel S, Bathla G, Murray J, Rhyner P. Preliminary Results from Retrospective Correlation of Circulating Tumor DNA (ct-DNA) with Imaging for HPV-Positive Oropharyngeal Squamous Cell Carcinoma. American Journal of Neuroradiology. Published online March 12, 2024. doi:10.3174/ajnr.A8242

17. Cao Y, Haring CT, Brummel C, Bhambhani C, Aryal M, Lee C, Heft Neal M, Bhangale A, Gu W, Casper K, Malloy K, Sun Y, Shuman A, Prince ME, Spector ME, Chinn S, Shah J, Schonewolf C, McHugh JB, Mills RE, Tewari M, Worden FP, Swiecicki PL, Mierzwa M, Brenner JC. Early HPV ctDNA Kinetics and Imaging Biomarkers Predict Therapeutic Response in p16+ Oropharyngeal Squamous Cell Carcinoma. Clinical Cancer Research. 2022;28(2):350–359. doi:10.1158/1078-0432.CCR-21-2338

18. Chotchutipan T, Rosen BS, Hawkins PG, Lee JY, Saripalli AL, Thakkar D, Eisbruch A, El Naqa I, Mierzwa ML. Volumetric ^18^ F FDG PET parameters as predictors of locoregional failure in low risk HPV related oropharyngeal cancer after definitive chemoradiation therapy. Head Neck. 2019;41(2):366–373. doi:10.1002/hed.25505

19. Fanizzi A, Comes MC, Bove S, Cavalera E, de Franco P, Di Rito A, Errico A, Lioce M, Pati F, Portaluri M, Saponaro C, Scognamillo G, Troiano I, Troiano M, Zito FA, Massafra R. Explainable prediction model for the human papillomavirus status in patients with oropharyngeal squamous cell carcinoma using CNN on CT images. Sci Rep. 2024;14(1):14276. doi:10.1038/s41598-024-65240-9

20. Kann BH, Likitlersuang J, Bontempi D, Ye Z, Aneja S, Bakst R, Kelly HR, Juliano AF, Payabvash S, Guenette JP, Uppaluri R, Margalit DN, Schoenfeld JD, Tishler RB, Haddad R, Aerts HJWL, Garcia JJ, Flamand Y, Subramaniam RM, Burtness BA, Ferris RL. Screening for extranodal extension in HPV-associated oropharyngeal carcinoma: evaluation of a CT-based deep learning algorithm in patient data from a multicentre, randomised de-escalation trial. Lancet Digit Health. 2023;5(6):e360–e369. doi:10.1016/S2589-7500(23)00046-8

21. Samarasinghe G, Jameson M, Vinod S, Field M, Dowling J, Sowmya A, Holloway L. Deep learning for segmentation in radiation therapy planning: a review. J Med Imaging Radiat Oncol. 2021;65(5):578–595. doi:10.1111/1754-9485.13286

22. Wong J, Huang V, Wells D, Giambattista J, Giambattista J, Kolbeck C, Otto K, Saibishkumar EP, Alexander A. Implementation of deep learning-based auto-segmentation for radiotherapy planning structures: a workflow study at two cancer centers. Radiation Oncology. 2021;16(1):101. doi:10.1186/s13014-021-01831-4

23. Lucido JJ, DeWees TA, Leavitt TR, Anand A, Beltran CJ, Brooke MD, Buroker JR, Foote RL, Foss OR, Gleason AM, Hodge TL, Hughes CO, Hunzeker AE, Laack NN, Lenz TK, Livne M, Morigami M, Moseley DJ, Undahl LM, Patel Y, Tryggestad EJ, Walker MZ, Zverovitch A, Patel SH. Validation of clinical acceptability of deep-learning-based automated segmentation of organs-at-risk for head-and-neck radiotherapy treatment planning. Front Oncol. 2023;13. doi:10.3389/fonc.2023.1137803

24. Wahid KA, Ahmed S, He R, van Dijk L V., Teuwen J, McDonald BA, Salama V, Mohamed ASR, Salzillo T, Dede C, Taku N, Lai SY, Fuller CD, Naser MA. Evaluation of deep learning-based multiparametric MRI oropharyngeal primary tumor auto-segmentation and investigation of input channel effects: Results from a prospective imaging registry. Clin Transl Radiat Oncol. 2022;32:6–14. doi:10.1016/j.ctro.2021.10.003

25. Cardenas CE, Anderson BM, Aristophanous M, Yang J, Rhee DJ, McCarroll RE, Mohamed ASR, Kamal M, Elgohari BA, Elhalawani HM, Fuller CD, Rao A, Garden AS, Court LE. Auto-delineation of oropharyngeal clinical target volumes using 3D convolutional neural networks. Phys Med Biol. 2018;63(21):215026. doi:10.1088/1361-6560/aae8a9

26. Reinders FCJ, Savenije MHF, de Ridder M, Maspero M, Doornaert PAH, Terhaard CHJ, Raaijmakers CPJ, Zakeri K, Lee NY, Aliotta E, Rangnekar A, Veeraraghavan H, Philippens MEP. Automatic segmentation for magnetic resonance imaging guided individual elective lymph node irradiation in head and neck cancer patients. Phys Imaging Radiat Oncol. 2024;32:100655. doi:10.1016/j.phro.2024.100655

27. Fakih M, Sandhu J, Wang C, Kim J, Chen YJ, Lai L, Melstrom K, Kaiser A. Evaluation of Comparative Surveillance Strategies of Circulating Tumor DNA, Imaging, and Carcinoembryonic Antigen Levels in Patients With Resected Colorectal Cancer. JAMA Netw Open. 2022;5(3):e221093. doi:10.1001/jamanetworkopen.2022.1093

28. Yousefi B, LaRiviere MJ, Cohen EA, Buckingham TH, Yee SS, Black TA, Chien AL, Noël P, Hwang WT, Katz SI, Aggarwal C, Thompson JC, Carpenter EL, Kontos D. Combining radiomic phenotypes of non-small cell lung cancer with liquid biopsy data may improve prediction of response to EGFR inhibitors. Sci Rep. 2021;11(1):9984. doi:10.1038/s41598-021-88239-y

29. Magbanua MJM, Li W, Wolf DM, Yau C, Hirst GL, Swigart LB, Newitt DC, Gibbs J, Delson AL, Kalashnikova E, Aleshin A, Zimmermann B, Chien AJ, Tripathy D, Esserman L, Hylton N, van ‘t Veer L. Circulating tumor DNA and magnetic resonance imaging to predict neoadjuvant chemotherapy response and recurrence risk. NPJ Breast Cancer. 2021;7(1):32. doi:10.1038/s41523-021-00239-3

30. Tran HT, Heeke S, Sujit S, Vokes N, Zhang J, Aminu M, Lam VK, Vaporciyan A, Swisher SG, Godoy MCB, Cascone T, Sepesi B, Gibbons DL, Wu J, Heymach JV. Circulating tumor DNA and radiological tumor volume identify patients at risk for relapse with resected, early-stage non-small-cell lung cancer. Annals of Oncology. 2024;35(2):183–189. doi:10.1016/j.annonc.2023.11.008

31. Liu S, Zhu Y, Chen Y, Wang Y, Zhang D, Zhang J, Wang Y, Zhang A, Hu Q, Liu A. Circulating Tumor DNA Combining with Imaging Analysis for Lesion Detection of Langerhans Cell Histiocytosis in Children. Children. 2024;11(12):1449. doi:10.3390/children11121449

